# Adenovirus-specific T cells in adults are frequent, cross-reactive to common childhood adenovirus infections and boosted by adenovirus-vectored vaccines

**DOI:** 10.1101/2024.04.25.24306332

**Authors:** Rookmini Mukhopadhyay, Arnold W. Lambisia, Jennifer P. Hoang, Benjamin J. Ravenhill, Charles N. Agoti, Benjamin A.C. Krishna, Charlotte J. Houldcroft

**Affiliations:** Department of Genetics, University of Cambridge, Cambridge, CB2 3EH, United Kingdom; Kenya Medical Research Institute-Wellcome Trust Research Programme, PO Box 230-80108, Kilifi, Kenya; Cambridge Institute for Medical Research, School of Clinical Medicine, University of Cambridge, Cambridge, CB2 0XY, United Kingdom; Department of Medicine, University of Cambridge, Cambridge, CB2 0QQ, United Kingdom

**Keywords:** Cellular immunity, Vaccination, Viral vectors, DNA virus

## Abstract

Human adenoviruses (HAdVs) cause diverse disease presentations as pathogens, and are also used as viral vectors for vaccines and gene therapy products. Preexisting adaptive immune responses to HAdV are known to influence symptom severity, viral clearance and the success of viral vectored products. Of note, approximately 50% of the UK’s adult population has received at least one dose of a chimpanzee adenovirus vectored SARS-CoV-2 vaccine (ChAdOx1) since January 2021.

We used FluoroSpot analysis to quantify the interferon gamma (IFNγ) and interleukin-2 (IL2) responses of healthy blood donors to HAdV species A, B, C, D and F and chimpanzee adenovirus Y25, related to HAdV species E. We find that cellular immune responses to multiple species of human adenovirus are ubiquitous among healthy adult blood donors, and that stimulating PBMC with whole hexon peptide libraries induces a significantly greater IFNγ and IL2 response than using selected peptide pools alone. We then compared the cellular immune responses of ChAdOx1 recipients and control donors using PBMC collected in 2021, and found that homotypic and heterotypic IFNγ responses were significantly boosted in ChAdOx1 recipients but not controls. Finally, we show that in PBMC derived from blood donors, IFNγ responses are made to both conserved and variable regions of the hexon protein.

Future vaccination campaigns using adenoviral vectored vaccines will need to account for the pre-existing exposure of recipients to both circulating HAdVs and vaccines such as ChAdOx1, which convey polyfunctional antiviral T cell responses to even low seroprevalence HAdV types.

## Introduction

Adenoviruses (AdVs) are non-enveloped, double-stranded DNA viruses with icosahedral capsids. Their capsids comprise three proteins: hexon, penton and fiber ^1^. More than 100 human adenoviruses (HAdVs) have been identified to date, which have been classified into seven species (A to G). The majority of primary HAdV infections occur during the first five years of life, and cause symptoms ranging from upper and lower respiratory tract infections and keratoconjunctivitis, to gastro-intestinal disease and fulminant infection ^2^. Currently there is no approved treatment for adenovirus infection, and no vaccine available for civilian use ^3,4^. Different adenoviral species are associated with different kinds of disease, and recombination in the hexon, penton and fiber genes of a given adenovirus can alter its tissue tropism and resulting symptom profile ^5^.

There has been significant interest in the immune response against HAdVs, largely driven by the problem of pre-existing HAdV immunity against HAdV-derived vectors, which suppresses the efficacy of immunisation ^6^. This is likely due to a biasing of immune responses to memory responses against the AdV backbone rather than *de novo* responses to the vaccine antigen. Previous studies have primarily focused on HAdV-C5. Passive neutralising antibody (nAb) transfer experiments in naïve mice dampened immune stimulation by HAdV-C5 vectors, but to a lesser extent than the dampening observed in pre-immune mice. This demonstrates that nAbs alone cannot account for all pre-existing immunity, implicating AdV-specific T cells in the AdV-induced immune response ^7^. Passive transfer of CD8^+^ T cells into naïve mice significantly decreased the immune response induced by HAdV-C5 vectors, highlighting their role in immune dampening ^7^. The role of T cells in adenoviral clearance is further illustrated by the success of adoptive T-cell therapy, where HAdV-infected patients are treated by transfusion of HAdV-specific T cells, with an overall 75% response rate in 63 HAdV-positive HSCT patients across 10 clinical trials ^8^. These data reinforce the importance of T cells in resolving HAdV infection in healthy and immunocompromised individuals and their potential role in dampening the immunisation efficacy of HAdV-derived vectors due to cross-reactivity.

Previous studies have suggested that HAdV-specific T cells are cross-reactive, capable of recognising a broad range of HAdVs. This is exemplified by the ability of HAdV-specific T cells to recognise chimpanzee-derived AdVs ^9^. Cross-reactivity arises from the ability of T cells to recognise conserved peptide regions, usually in the hexon ^10^. MHC-II-restricted CD4^+^ epitopes have been identified throughout the hexon, but predominantly in the conserved regions ^9–11^, while MHC-I-restricted CD8^+^ T cell epitopes have also been identified mainly in the hexon, but also in the penton and fibre ^10,12,13^. T cell responses to conserved regions are presumably recalled repeatedly upon infection with different HAdVs, whereas responses to variable regions are less frequently stimulated ^9^. By contrast, nAbs show limited cross-neutralisation of different HAdV subtypes, which is a consequence of nAbs targeting peptide regions with high heterogeneity, such as the hexon hypervariable regions (HVRs) ^14–16^.

In addition to the cellular immune landscape of natural HAdV infection, approximately 50% of the UK’s adult population have received ChAdOx1, the Oxford-Astra Zeneca SARS-CoV-2 vaccine ^17^. This is a viral-vectored vaccine which is based on chimpanzee adenovirus Y25 (GenBank accession: JN254802), and contains a deletion of Y25 genes E4, ORFs 4, 6, 7 and the 34K CDS region and insertion of the equivalent portion of the human adenovirus C5 genome ^18,19^.

We therefore set out to investigate the interferon gamma (IFNγ) and interleukin 2 (IL2) T cell response of healthy blood donors to diverse human adenoviruses from multiple species; to establish whether the T cell response to a non-species C human adenovirus was confined to the conserved regions of the hexon protein; and to quantify what effect the use of ChAdOx1 has had on the landscape of anti-adenovirus T cell immunity in UK donors.

## Methods

### Donors

#### Healthy blood donors

Samples from ten anonymised healthy blood donors were collected from NHS Blood and Transplant (NHSBT), Cambridge Donor Centre. Ethical permission for “Understanding humoral and cellular immune responses to DNA viruses in healthy blood donors” was granted by the HRA and Health and Care Research Wales (HCRW) (REC reference 22/WA/0162). PBMC and serum were collected from leukocyte reduction system cones, a by-product of the platelet donation process. PBMC were separated from cones following a previously published protocol ^20^ using pluriSelect PBMC 24+ Spin Medium (Cambridge Bioscience, Cambridge, UK). Platelet donors from whom leukocyte cones were derived are aged between 17 and 70; specific age and sex data for individual donors was not available.

Twenty one donors of known SARS-CoV-2 vaccine status were recruited in 2021, all patients gave informed written consent in accordance with the Declaration of Helsinki. Ethical permission for the ARIA (Anti-viral Responses in Ageing, CBR53) study was granted by the Cambridge Human Biology Research Ethics Committee (HBREC.2014.07). Donors were grouped as recipients of one or more doses of ChAdOx1 vaccine (ChAdOx1 recipients), n = 11; or one or more doses of mRNA vaccine or no vaccine at the point of blood donation (controls), n = 10.

For both cohorts, PBMC were separated, frozen and thawed as previously described ^21^. Cell viability was determined using trypan blue exclusion staining and counting of live cells using a haemocytometer.

### Serology

In blood donors, IgG responses to HAdV-C5 were measured by ELISA using a Human Adenovirus IgG (ADV-IgG) ELISA Kit [AE24150HU] (Abebio, Wuhan, China), following the manufacturer’s recommended protocol. For samples 2301-2304, haemolysate was used to counter erythrocyte contamination due to NHSBT cone storage duration of longer than 12 hours before serum sampling; for samples 2305-2310, serum was used.

### Peptide stimulants

#### Adenovirus ORF and other peptide mixes

Six commercially available, and two custom, peptide pools were selected to represent the diversity of human adenovirus species (**Table 1**). Commercial peptide pools from Miltenyi and JPT (Table 1) were diluted to a concentration of 5μg/ml/peptide. A custom library of consecutive 15-mer peptides overlapping by 5 amino acids were synthesised by GenScript (Oxford, UK) using the HAdV-A12 hexon sequence (GenBank accession: NP_040924.1). Individual lyophilised peptides from each custom ORF library were reconstituted in 20% DMSO-80% RPMI-1640 (Sigma) at 10mg/ml master stock. Individual peptides were then diluted in RPMI-1640 to give a 1mg/ml (2% DMSO) working stock. Peptide pools were used as either entire ORF mixes at a concentration of 5μg/ml/peptide (final working concentration shown in Table 1) or formed into pools of 40-60 peptide pool of conserved and variable epitopes (see below), at a concentration of 20μg/ml/peptide.

**Table 1:**

| Name | Supplier | Adenovirus species and type | Final concentration |
| --- | --- | --- | --- |
| Custom HAdV A12 hexon pool | GenScript | A: 12 | 2µg/ml/peptide |
| HAdV-A conserved and variable pools | GenScript | A: 12 | 20µg/ml/peptide |
| PepMix HAdV-3 hexon (PM-HAdV3) | JPT | B: 3 | 2µg/ml/peptide |
| PepTivator AdV Select (130-124-394)* | Miltenyi | C: 2 and 5 | 2µg/ml/peptide |
| PepTivator AdV5 Hexon (130-093-495) | Miltenyi | C: 5 | 2µg/ml/peptide |
| PepTivator AdV5 Penton (130-096-777) | Miltenyi | C: 5 | 2µg/ml/peptide |
| PepMix Human Adenovirus 26 Hexon (PM-HADV26-L3-1) | JPT | D: 26 | 2µg/ml/peptide |
| PepMix Human Adenovirus 26 Penton (PM-HADV26-L2-1) | JPT | D: 26 | 2µg/ml/peptide |
| Custom HAdV F41 hexon pool | GenScript | F: 41 | 2µg/ml/peptide |
| PepMix Chimpanzee Adenovirus Y25 Hexon (PM-CHADVY25-L3-1) | JPT | ChAd: Y25 | 2µg/ml/peptide |
| PepMix Chimpanzee Adenovirus Y25 Penton (PM-CHADVY25-L2-1) | JPT | ChAd: Y25 | 2µg/ml/peptide |

\*AdV Select is a pool of 50 MHC class I and class II restricted oligopeptides derived from human adenovirus species C genotypes 2 and 5.

#### Defining conserved versus variable regions

Peptides were defined as conserved if there were eight consecutive amino acids out of fifteen identical between the C5 and A12 hexon amino acid reference sequences (AP_000211.1 and NP_040924.1 respectively). The epitopes constituting the A12 conserved 1, conserved 2 and variable pools are listed in **SUPPLEMENTARY TABLE 1.**

### Detection of Cytokine Production in PBMC by FluoroSpot

PBMC were incubated in pre-coated human IFNγ and IL2 FluoroSpot plates (Mabtech AB, Nacka Strand, Sweden) in triplicate with ORF peptide pools (final peptide concentration shown in **TABLE 1** following dilution with TexMacs) and an unstimulated and positive control mix [containing anti-CD3 and anti-CD28 (ImmunoCult Human CD3/CD28 T Cell Activator, StemCell)], for 48h at 37°C. Cells and media were decanted from the plate and developed following the manufacturer’s protocol. After development and drying overnight, plates were read using an AID iSpot reader (Oxford Biosystems, Oxford, UK) and spot-forming units were counted using AID EliSpot v7 software (Autoimmun Diagnostika GmbH, Strasberg, Germany).

The mean spot forming units (SFU) were converted to SFU per 10^6^ PBMC, the mean background response (SFU/10^6^ cells) was deducted from the mean of the corresponding wells. The cutoff for a positive response was 50 SFU/10^6^ cells for IFNγ responses and 10 SFU/10^6^ for IL2 responses, defined based on responses to SARS-CoV-2 and adenovirus vectored vaccines as no clinical T cell correlate of protection is currently defined for human adenovirses ^22–24^. Values at or below zero were plotted as 0.1 to allow their visualisation on logarithmic axes.

Donors were excluded from further analysis if they failed to produce above-background IFNγ responses to positive control stimulation, compared to the negative control.

### Multiple sequence alignment and amino acid distance calculations

Hexon amino acid sequences for HAdVs A12 (NP_040924.1), C5 (AP_000211.1), B3 (YP_002213779.1), D26 (ABO61316.1), F41 (ACH90432.1) and ChAd Y25 (YP_006272963.1) were retrieved from GenBank. Sequences were aligned in MEGA11 using MUSCLE and manually inspected. Estimated evolutionary distances between amino acid sequences were calculated in MEGA11 as the number of amino acid substitutions per site between sequences, with a Poisson correction model. Ambiguous positions were removed for each sequence pair using the pairwise deletion option. There were 976 positions in the final dataset.

### Statistical analyses

Data were tested for normality using Shapiro–Wilk tests and parametric or non-parametric statistical tests chosen accordingly. Spearman’s R was calculated for the averaged background corrected SFU per 10^6 PBMC for each pair of hexon proteins. SFU which were 0 after background correction are represented as 0.1 on log scale graphs. Unless otherwise stated, statistical analysis and graphing were performed in GraphPad Prism v10.2. A Mantel test of the relationship between hexon pairwise amino acid distances and IFNγ response correlations was performed in Matlab 2021b, using the Fathom toolbox (Jones, 2017), set at 9999 permutations.

## Results

### Serological responses to adenovirus C5 are relatively uncommon

Serum antibody responses to adenovirus are often used as proxy for previous infection history ^26^. We therefore measured anti-adenovirus C5-IgG levels by ELISA in each of our healthy donors. 2/10 donors had a positive IgG response to adenovirus C5, and 4/10 had equivocal responses (confidence intervals overlapping the cutoff). The remaining four donors were seronegative. This is broadly in line with binding antibody data collected in Germany ^27,28^, where binding antibody levels were higher for genotype C1 than C5, and for previous serology surveys conducted in Europe ^26^. This may also mirror the significant post-COVID reduction in binding antibody levels seen in German healthy serum donors between 2019 and 2021 ^28^, but the numbers presented here are small. No comparable data is available for England.

### Interferon gamma-producing responses to adenoviruses are pervasive and IL2 responses are seen in the majority of healthy donors

IFNγ responses, measured by FluoroSpot or similar technologies, are an *in vitro* proxy for *in vivo* antiviral T cell recognition and response to viral peptides. T cell responses are an alternative tool for identifying previous viral pathogen exposure in seronegative individuals ^22^. Using peptides derived from prevalent adenovirus species C genotypes, we found that 7/10 donors made a positive IFNγ response to the AdV Select (C2 and C5) pool, and 8/10 to the C5 hexon peptide pool. The AdV Select pool is a set of peptides spanning epitopes experimentally identified from genotypes 2 and 5, while the C5 hexon pool tiles the whole of the immunodominant hexon protein. All donors made a positive IFNγ response to the hexons of HAdVs-A12, B3, and D26, demonstrating clear exposure to a range of HAdV species even in the absence of detectable antibody serology.

IL2 responses are a proxy for CD4^+^ T cell recognition of viral peptides ^29^. Similar to IFNγ, 7/10 donors had a positive IL2 response to the C5 hexon pool and 3/10 donors had a positive response to the C5 penton pool. However, fewer donors made a positive response IL2 (4/10 donors) to the AdV Select pool, compared to IFNγ (7/10).

### Interferon gamma and interleukin-2 responses to adenovirus penton peptide pools are less common in healthy blood donors than hexon responses

The adenovirus penton protein is a known to be a target of human T cell responses, though the relative contribution of the penton and hexon to the T cell response to adenovirus infection is currently an area of active research ^30^. We therefore quantified the IFNγ and IL2 responses to two commercially available human adenovirus penton peptide pools: HAdVs-C5 and D26.

We found that positive IFNγ responses to the C5 penton were less common than C5 hexon responses, with 3/10 donors responding to the C5 penton peptide pool (**Figure 3**) compared to 8/10 responding to the hexon pool (**Figure 2**). The same was also true of D26 (5/10 responding to the penton, compared to 10/10 responding to the hexon).

**Figure 1 legend:**
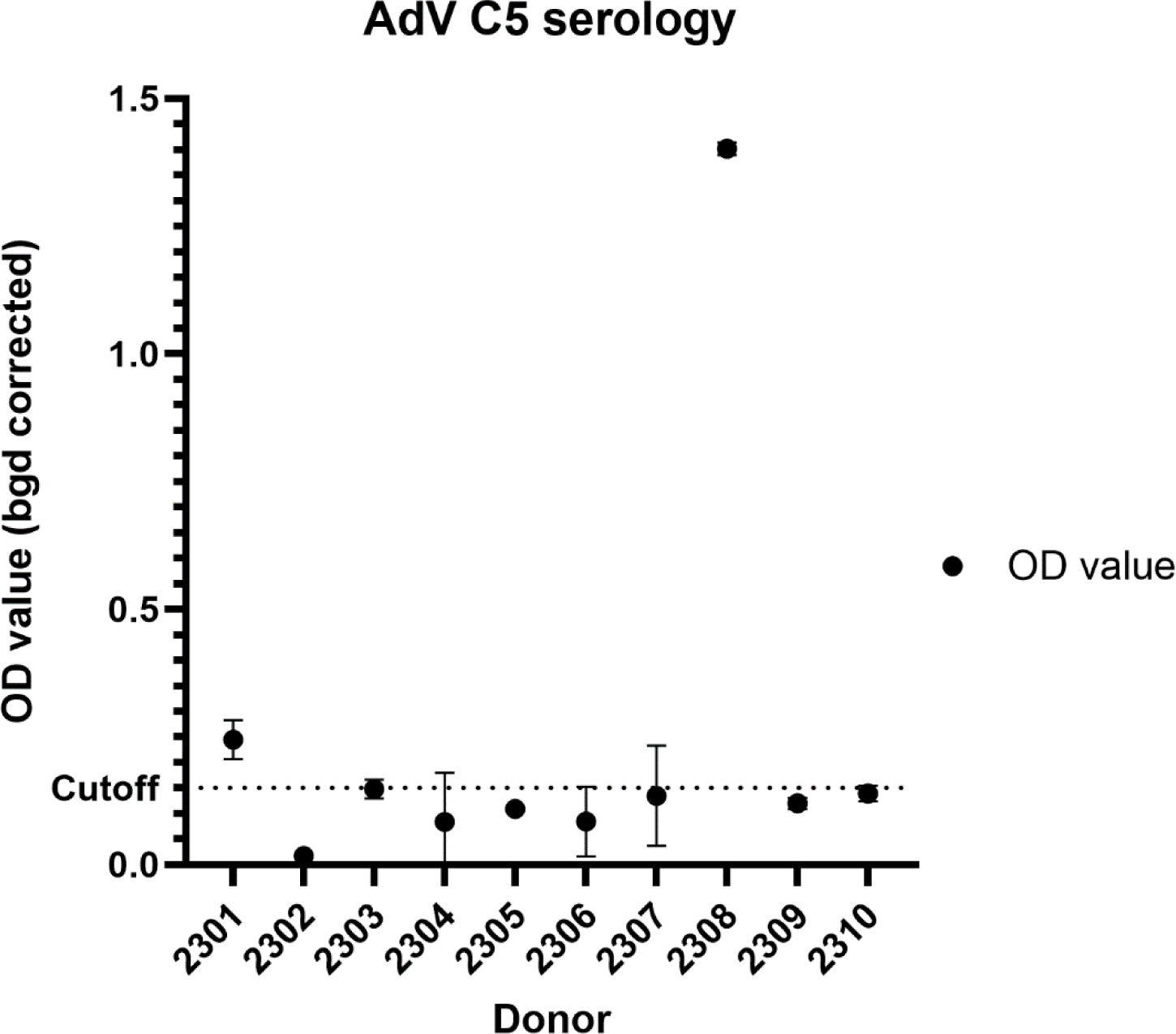
Plot showing serum antibody responses to adenovirus C5 in healthy blood donors. Optical density measurements were made using an ELISA specific for HAdV C5 and were background-corrected.

**Figure 2 legend:**
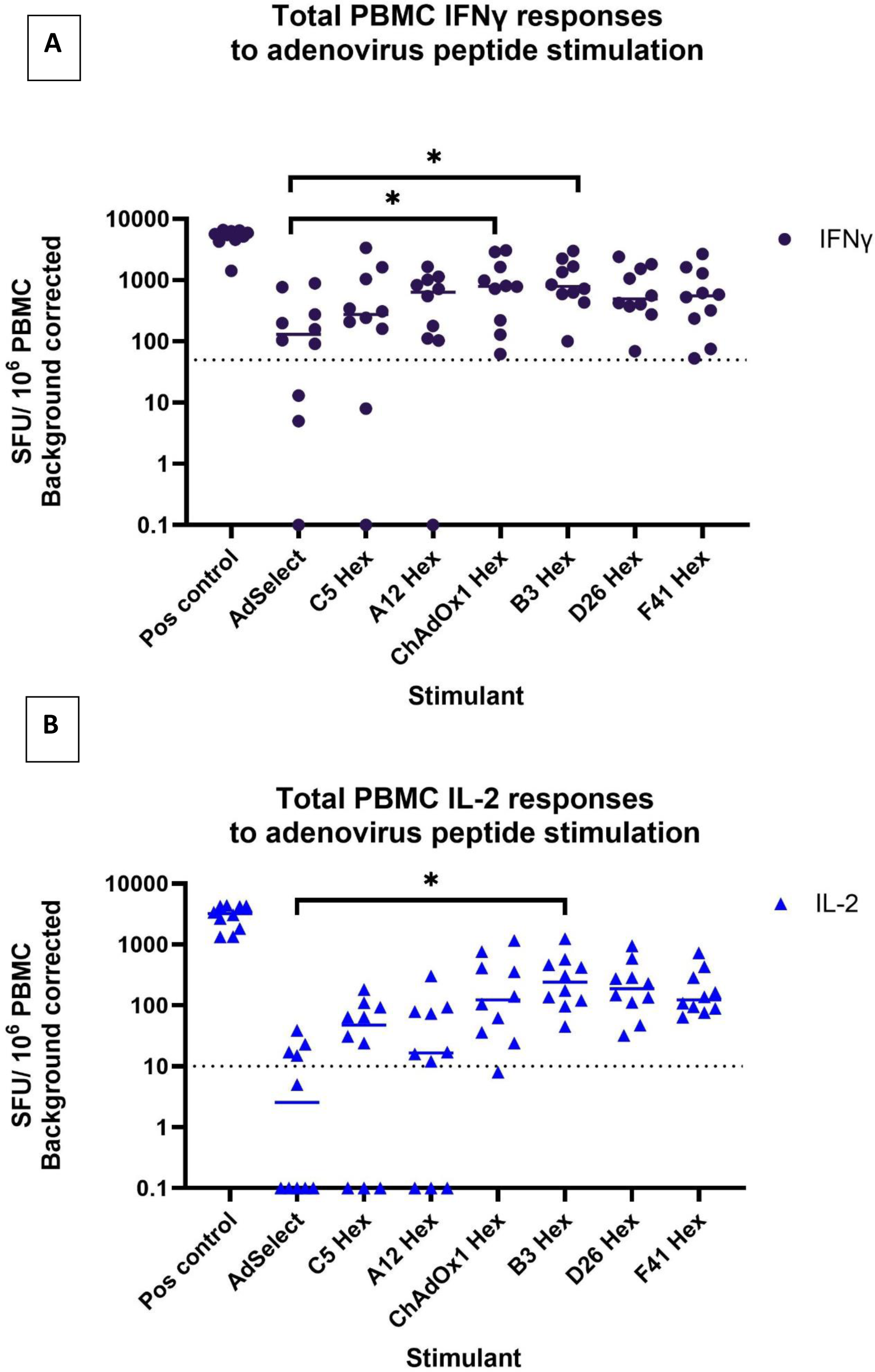
Analysis of AdV specific IFNγ and IL2 FluoroSpot responses to AdV peptide pools (A) IFNγ FluoroSpot responses to AdV peptide pools covering: an experimentally-validated set of 50 AdV C2 and 5 peptides (AdSelect), peptide pools covering the entire hexons of human adenoviruses A12, B3, C5, D26, F41 and the hexon of chimpanzee adenovirus Y25 (vector backbone of SARS-CoV-2 vaccine ChAdOx1), as well as a polyclonal anti-CD3/CD28 antibody T cell stimulation as a positive control of PBMC from healthy blood donors, calculated as spot-forming units (SFU) per 10e6 PBMC (background corrected). (B) IL2 FluoroSpot responses to AdV peptide pools covering: an experimentally-validated set of 50 AdV C2 and 5 peptides (AdSelect), pools covering the entire hexons of human adenoviruses A12, B3, C5, D26, F41, the hexon of chimpanzee adenovirus Y25 (vector backbone of SARS-CoV-2 vaccine ChAdOx1), as well as a polyclonal anti-CD3/CD28 antibody T cell stimulation as a positive control of PBMC from healthy blood donors, calculated as spot-forming units (SFU) per 10e6 PBMC (background corrected). Significance determined by two-way ANOVA, corrected for multiple testing. Key: *p < 0.05. The dotted lines indicate the boundary between a positive and a negative response.

**Figure 3 legend:**
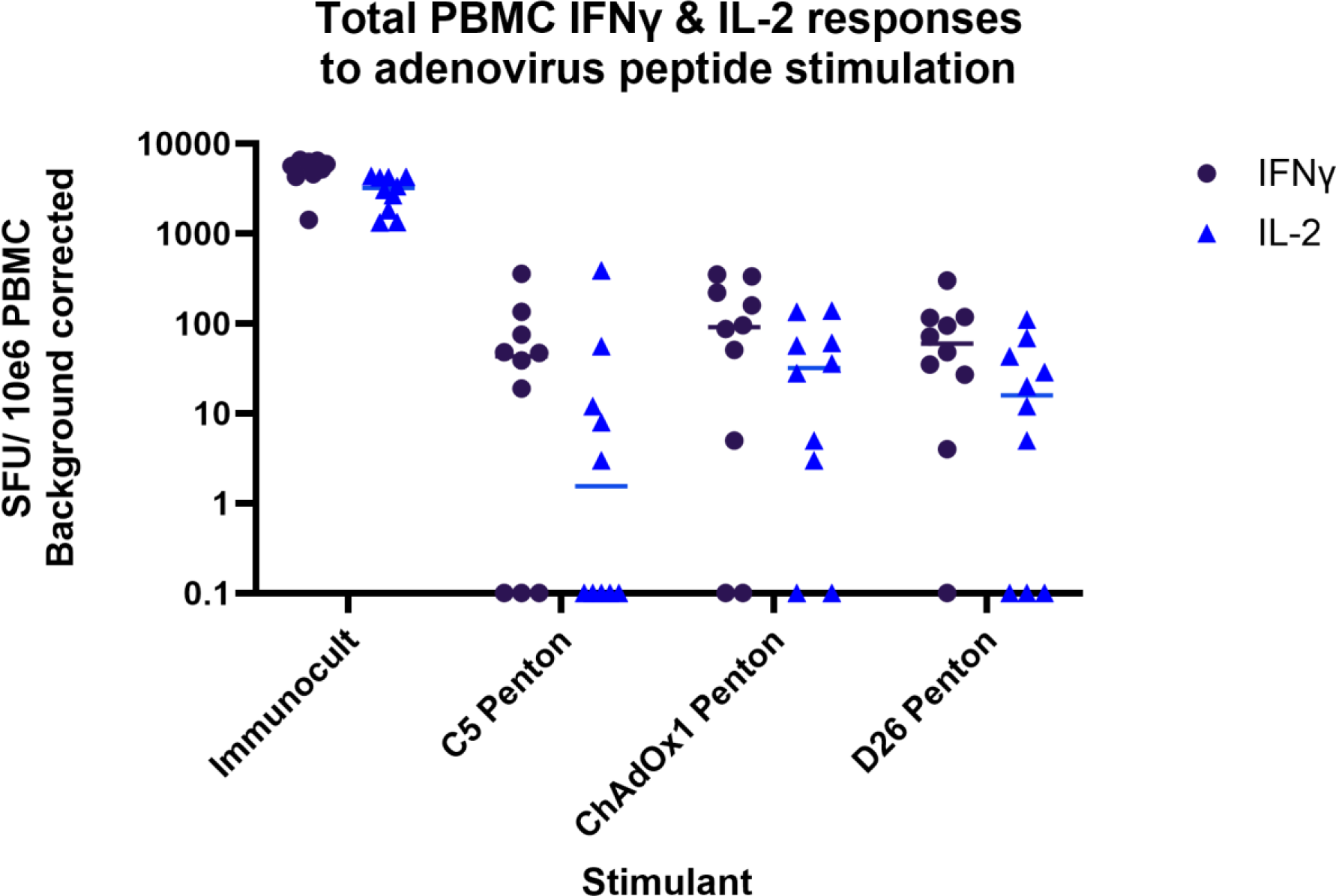
Analysis of AdV specific IFNγ and IL2 FluoroSpot responses to AdV peptide pools (A) IFNγ FluoroSpot responses to AdV peptide pools covering: the pentons of human adenoviruses C5 and D26, and the penton of chimpanzee adenovirus Y25 (vector backbone ChAdOx1) as well as a polyclonal anti-CD3/CD28 antibody T cell stimulation as a positive control of PBMC from healthy blood donors, calculated as spot-forming units (SFU) per 10e6 PBMC (background corrected). (B) IL2 FluoroSpot responses to AdV peptide pools covering: the pentons of human adenoviruses C5 and D26, and the penton of chimpanzee adenovirus Y25 (vector backbone ChAdOx1) as well as a polyclonal anti-CD3/CD28 antibody T cell stimulation as a positive control of PBMC from healthy blood donors, calculated as spot-forming units (SFU) per 10^6 PBMC (background corrected).

Similar trends were seen for IL2 responses, with 2/10 donors responding to penton peptide pools derived from C5, and 6/10 responding to D25 (**Figure 3**).

### IFNγ and IL2 responses to the hexon of the ChAdOx1 vector, chimpanzee adenovirus Y25, are ubiquitous among healthy blood donors recruited in 2023

In 2021-2022, an estimated 50% of the UK’s adult population received at least one dose of an adenovirus-vectored SARS-CoV-2 vaccine, predominantly ChAdOx1 ^17^. The ChAdOx1 vector is based on ChAd-Y25, retaining the ChAd-Y25 hexon and penton ^18,19^. ChAd-Y25 has homology to HAdV species E ^19^, which is associated with outbreaks of respiratory disease in congregate settings ^31^.

We found that 10/10 healthy blood donors made a positive IFNγ response to the hexon of ChAd-Y25 (ChAdOx1) and 9/10 donors made a positive IL2 response (**Figure 2**). Similar to observations of human adenovirus-derived penton peptide pools, fewer healthy donors made a positive IFNγ (7/10) or IL2 (5/10) response to the ChAdOx1 penton (**Figure 3**) than the hexon.

### Positive IFNγ responses to ChAdOx1 and C5 hexon peptide pools are more common in ChAdOx1 vaccine recipients than controls

Among anonymised apheresis cones from healthy blood donors recruited in 2023, ChAdOx1 vaccination status was not available. We hypothesised that among healthy donors of known ChAdOx1 recipient status, ChAdOx1 recipients would have higher T cell responses to the ChAdOx1 vector hexon than recipients who had received an mRNA SARS-CoV-2 vaccine or no vaccine instead. We also hypothesised that boosting of C5 hexon T cell responses might occur due to vaccine-induced activation of cross-reactive T cells which recognised shared epitopes. We therefore compared PBMC responses to the ChAdOx1 and C5 hexons in healthy controls and ChAdOx1 recipients of known vaccine status, vaccinated and recruited in 2021.

Positive IFNγ responses to ChAdOx1 and C5 hexon peptide pools, defined as >50 SFU/10e6 cells, were more common among ChAdOx1 SARS-CoV-2 vaccine recipients than controls (mRNA SARS-CoV-2 vaccine or no vaccine). 7/10 ChAdOx1 recipients had a positive IFNγ response to the ChAdOx1 hexon peptide pool while 2/11 controls had a positive response to Y25. 5/10 ChAdOx1 recipients had a positive IFNγ response and 1/11 controls had a positive IFNγ response to the C5 peptide pool. We found that IFNγ T cell responses were statistically significantly higher in ChAdOx1 recipients for both their responses to the Y25 (ChAdOx1) hexon (mean SFU/10e6 cells 143.1, SD 169.1 vs mean SFU/10e6 cells 79.6, SD 203.7; Mann Whitney U test (two-tailed) p = 0.023) and the C5 hexon (mean SFU/10e6 cells 66.01, SD 49.45 vs mean SFU/10e6 cells 18.8, SD 48.5; Mann Whitney U test (two tailed) p = 0.005) peptide pools.

IL2 responses were similar between ChAdOx1 recipients and controls. 5/10 ChAdOx1 recipients had a positive IL2 response to the Y25 (ChAdOx1) hexon peptide pool, defined as >10 SFU/10e6 cells, while 1/11 controls had a positive response. 4/10 ChAdOx1 recipients and 2/11 controls had a positive IL2 response to the C5 hexon peptide pool. The differences in mean SFU between the two groups were not statistically significant (Mann Whitney U test (two tailed), Y25 p = 0.19; C5 p = 0.34).

There was a statistically significant correlation in the frequency of IFNγ T cell responses to Y25 and C5 in the 2021 vaccine recipients (r2 = 0.87, p = <0.0001) (**supplementary figure 1**). This appears to be driven largely by the ChAdOx1 recipients (r2 = 0.82, p = 0.0001), as the correlation was not statistically significant in the controls (R2 = 0.03, p = 0.6). There was no statistically significant correlation between the frequency of IL2 T cell responses to Y25 and C5 in the 2021 vaccine recipients (r2 = 0.06, p = 0.42; data not shown) or any subgroup.

### Relationship between the frequency of cytokine responses between species

Adenovirus-specific T cell responses have previously been reported to be cross-reactive within and between adenovirus species ^9,32^. Therefore, we hypothesised that cross-reactive T cells from the same donor should recognise peptides from multiple different adenovirus species, and that the magnitude of the response should be positively correlated: i.e. the IFNγ and IL2 response should be produced by a mixture of type or species-specific T cells, and also a population of cross-reactive T cells. Previous research suggests that the proportion of cross-reactive T cells should be higher than the proportion of type/species-specific T cells.

We investigated this by calculating the correlations in the magnitude of SFU responses between each pair of hexon sequences. A number of pairs of hexon sequences had statistically significant Spearman’s r coefficients for the frequency of IFNγ responses (**FIGURE 5**). There was a statistically significant relationship between the IFNγ and IL2 responses for genotypes A12 and ChAdOx1 (Y25), B3 and D26, B3 and F41, and D26 and F41. IL2 response correlations are shown in **supplementary figure 2**. Correlations between C5 and D26 and C5 and F41 were only significant for IFNγ responses.

Not all pairs of hexons had significantly correlated IFNγ and/or IL2 response frequencies. This suggests that cross-reactivity of adenovirus-specific T cell responses may not apply equally across adenovirus species. Furthermore, a Mantel test of the correlation between genetic distances of paired hexon amino acid sequences, and the correlations in IFNγ responses between pairs of hexons, was not statistically significant. This suggests that shared infection history may explain the correlations between hexon pairs, rather than the degree of amino acid conservation.

### Many donors make IFNγ T cell responses to variable regions of the hexon

In adenovirus C, previous work had identified conserved regions of the hexon protein to be important for the T cell response, and in particular the CD4^+^ T cell response (eg ^33^). In order to understand the relative contribution of variable and conserved regions of the hexon to the T cell response to adenovirus, the adenovirus A12 hexon was divided into three pools: one containing variable epitopes which are not highly conserved between adenovirus species, and two conserved pools, a 5’ conserved pool 1 and a 3’ conserved pool 2 (**supplementary table 1**) (**Figure 6A**). The variable pool included peptides which showed variation within species A genotypes. In a post-SARS-CoV-2 emergence sample of donor serum from Germany, binding antibody responses to A12 were higher on average than responses to other species A genotypes, suggesting A12 was a suitable and seroprevalent representative of adaptive immune responses to species A adenoviruses in a post-pandemic population ^27^.

We found the cellular immune response to A12 to include detectable IFNγ responses to all regions of the protein, while the IL2 responses was more variable from donor to donor. 5/6 healthy blood donors made an IFNγ response to the variable portion of the hexon, while all donors made an IFNγ response to the 5’ and 3’ conserved regions. The IL2 response to the variable domain was absent in 4/6 donors, in contrast to the IFNγ response. 5/6 donors made an IL2 response to 5’ conserved pool 1, and 3/6 made responses to 3’ conserved pool 2. This may reflect a bias of CD4^+^ T cell responses towards the more conserved regions of the hexon protein, which is less marked for CD8^+^ T cells.

## Discussion

The *de novo* T cell response to adenovirus infection is widely recognised as ameliorating the severity of disease ^34^, while also playing a role in the success of adenovirus-vectored vaccines and gene therapy products ^35^. In this study, we investigated the frequency and function of adenovirus-specific T cell responses in healthy donors using highly sensitive FluoroSpot assays, comparing responses to adenovirus proteins derived from five human and one chimpanzee adenovirus species. We also compared the effect of an adenovirus-vectored vaccine (ChAdOx1) on T cell responses to the vector (ChAdV-Y25) and a commonly used human adenovirus vector (HAdV-C5).

We note a significant dichotomy between the high frequency of T cell responses to different adenovirus species (Figure 2), and the relatively small frequency of donors with detectable binding antibody levels to HAdV-C5 (Figure 1). Recent data from healthy donors in Germany ^27,28^ suggests that this may be because that the majority of donors have low-level circulating binding antibody responses to C5, represented by low OD values. We speculate that ELISA-based serological analysis of adenovirus binding antibody responses may not be sufficiently sensitive to establish past adenovirus infection history and immunity on a per-genotype level. As with SARS-CoV-2, seroreversion may be a feature of infrequent adenovirus re-exposure and/or reinfection in the adult population ^22^. Additionally, the “hit and run” strategy seen in these respiratory viruses may lead to low levels of serum antibodies in favour of mucosal responses dominated by IgA.

The commercial Peptivator AdV Select pool of peptides (Miltenyi) has been used for the generation of therapeutic anti-HAdV T cell products by a number of groups eg ^36,37^, and consists of a defined, experimentally-validated set of HLA class I and II epitopes from HAdVs C2 and C5. We note that among healthy blood donors, more individuals made a detectable IL2 response to the HAdV C5 hexon pool than the AdV Select pool. The HAdV5 hexon peptide pool tiles the entire protein, and thus is agnostic to the HLA type of the donor. Indeed, both IFNγ and IL2 responses were statistically significantly more frequent to a peptide pool derived from the hexon of common respiratory HAdV B3 than to the AdV Select pool (**Figure 2**). This suggests that in donors of unknown HLA type, using a HAdV hexon pool which tiles the entire protein leads to more donors having a detectable IFNγ and IL2 T cell response than the AdV Select pool alone when generating therapeutic T cells for AdV cellular therapy.

25 million first doses of the ChAdOx1 adenovirus-vectored SARS-CoV-2 vaccine were administered to the UK’s adult population received between 2021 and 2023 ^17^. Y25 is the vector backbone from which ChAdOx1 was derived. In 2012, neutralising antibody responses to the vector were reported to be low in the UK population, with no donors having neutralising antibody titres where ND50 is at serum dilutions over 200 ^19^; after immunisation with one or more doses of ChAdOx1, anti-vector IgG responses became detectable ^38^. Other studies have also identified that use of a HAdV-C5 vectored COVID vaccine boosts neutralising antibody responses in seronegative recipients ^39^. To the best of our knowledge, this is the first study to explore the degree of anti-vector T cell responses to Y25 since the UK’s ChAdOx1 vaccination campaign. Among platelet donors at NHSBT Cambridge, from whom leukocyte reduction cones are derived, T cell responses to Y25 viral surface proteins are now ubiquitous and of comparable magnitude to common genotypes such as C5. In samples collected in 2021 from healthy donors, ChAdOx1 vaccine recipients had statistically significantly higher IFNγ responses to both Y25 and C5 hexons than controls, suggesting both a type-specific and cross-species boosting of cellular immunity to adenoviruses (**Figure 4, supplementary figure 1**). This may have important consequences for future vaccination campaigns or gene-therapy products wishing to use adenovirus vectors in individuals who have also received ChAdOx1.

**Figure 4 legend:**
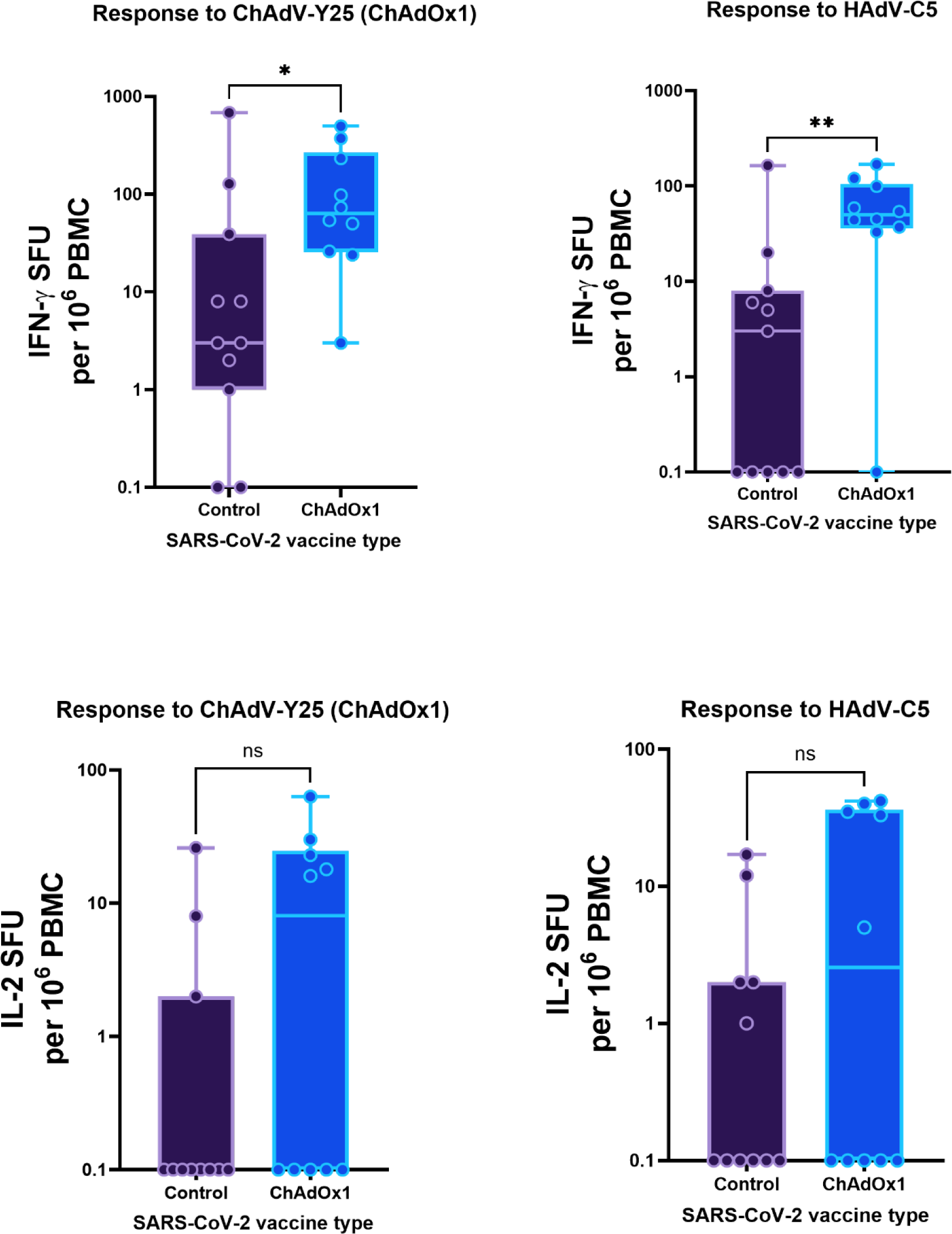
Frequency of IFNγ and IL2 responses to adenovirus peptide pools covering the hexon proteins of Y25 and C5 in ChAdOx1 vaccine recipients and controls, expressed as spot-forming units per 10e6 PBMC (background corrected). P values were calculated with a Mann-Whitney U test.

**Figure 5 legend:**
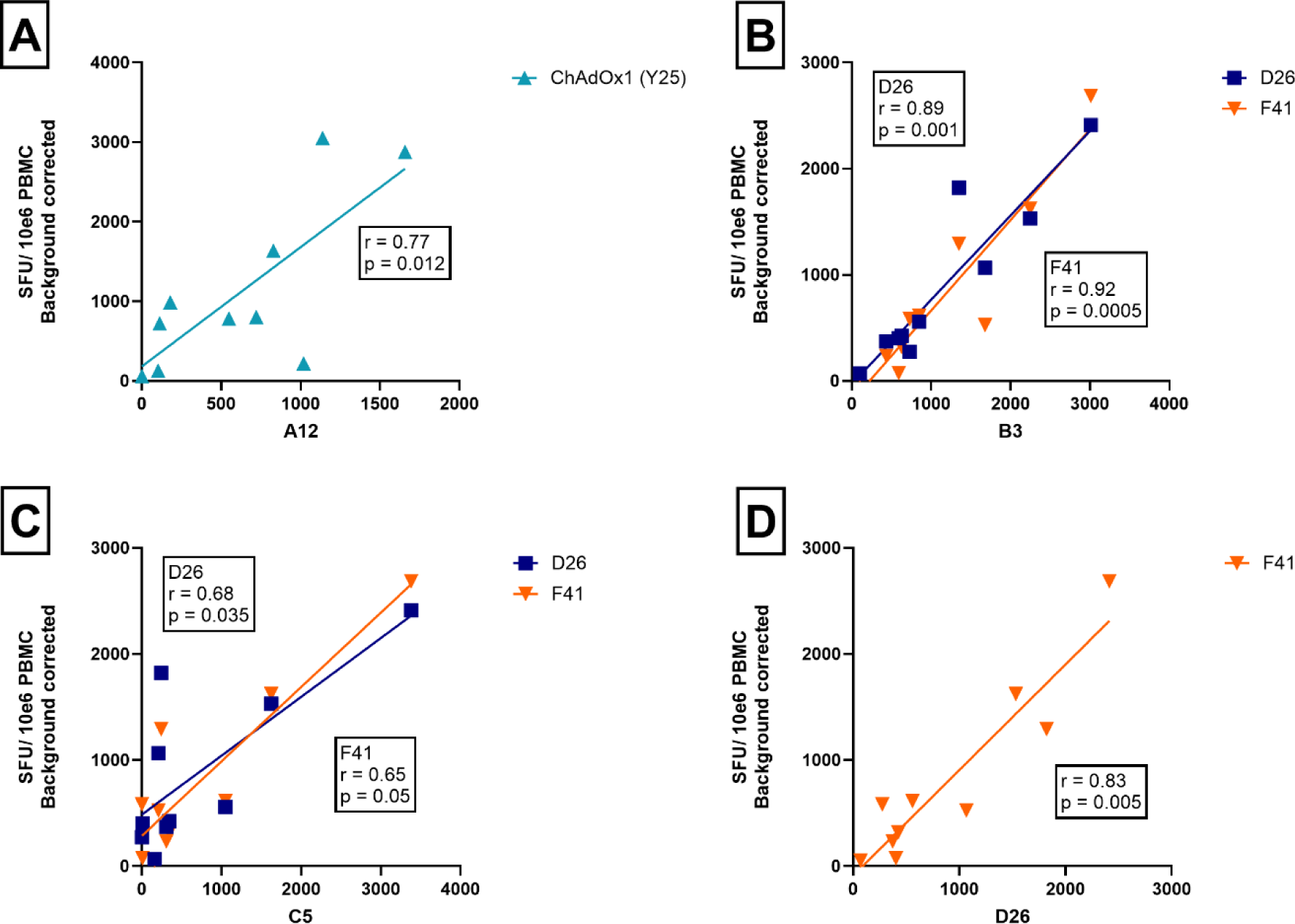
Plots showing the correlation between the frequency of IFNγ responses for pairs of hexons for ten healthy donors. Each axis shows the number of SFUs per 10^6^ PBMC produced by each donor in response to hexon peptide pool stimulation. Spearman’s r correlation coefficients and two-tailed p values shown for statistically significant correlations.

Previous studies have shown that the hexon T cell response is equally distributed against both the variable and conserved regions of the protein ^9^ while others have suggested it is focused on conserved epitopes ^10,33^. Our data suggest that IFNγ responses, a proxy for CD8^+^ T cell responses ^29^, may be more skewed towards variable regions than previously thought (**Figure 6B**). The weak or absent correlation in the frequency of T cell responses to some pairs of hexons of different adenovirus species, and the common recognition of variable epitopes within the A12 hexon peptide pool by healthy blood donor PBMC, suggests an important species- or genotype-specific component of the IFNγ response to adenovirus, which is therefore unlikely to be to conserved epitopes. There was no statistically significant correlation between amino acid distance between hexon pairs and the correlation in response frequency within donors, which suggests that shared infection history may explain the correlations in IFNγ responses to the genotypes of some hexon pairs. In contrast, IL2 responses (a proxy for the CD4^+^ T cell response ^40^) to the variable domain of the A12 hexon peptide pool were relatively unusual (2/6 donors), which supports previous studies on the CD4^+^ T cell response being focused towards conserved epitopes (**Figure 6C**). The relative contribution of different classes of T cell response to adenovirus infection in adults merits further research in order to refine future vaccination and cellular therapy efforts.

**Figure 6 legend:**
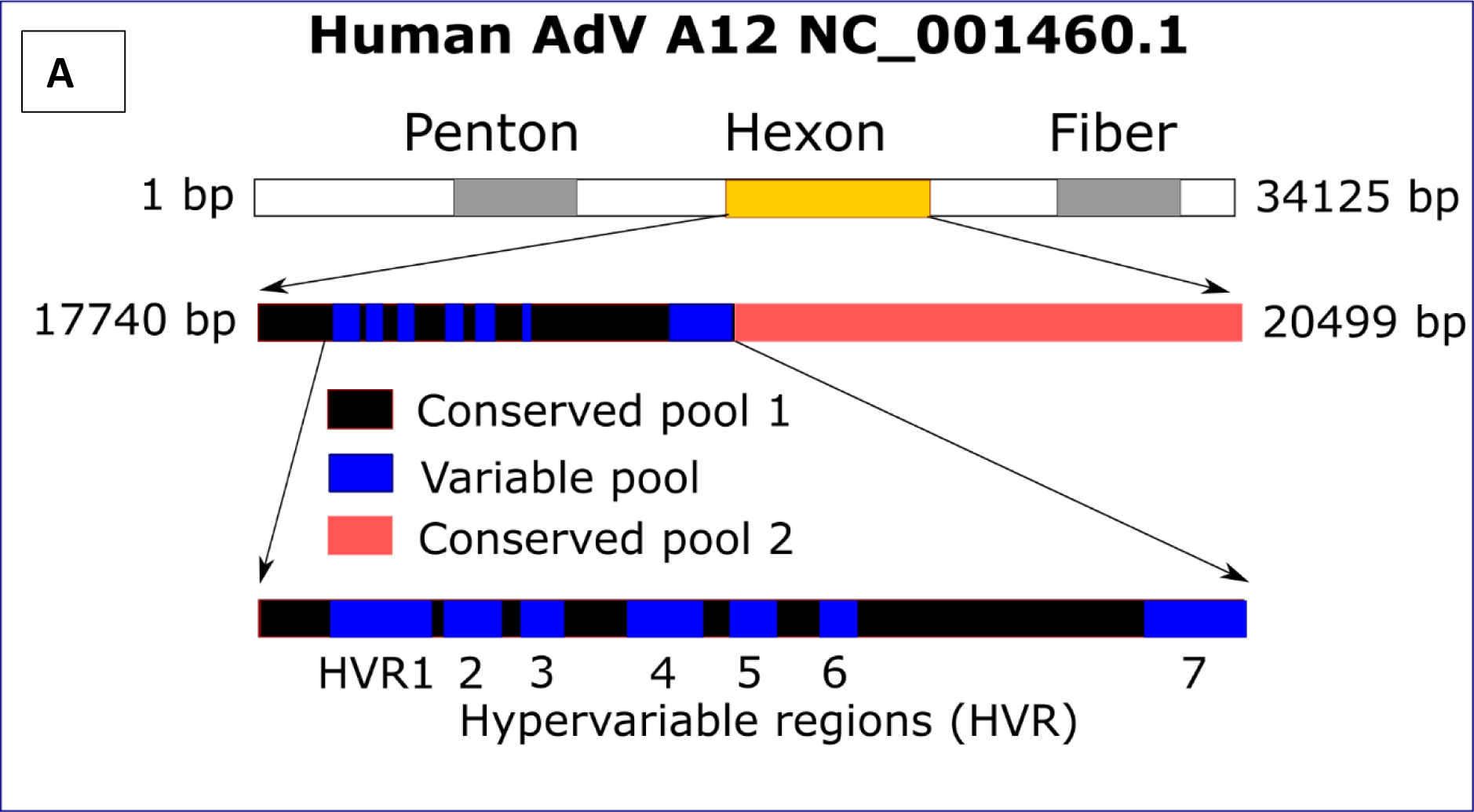
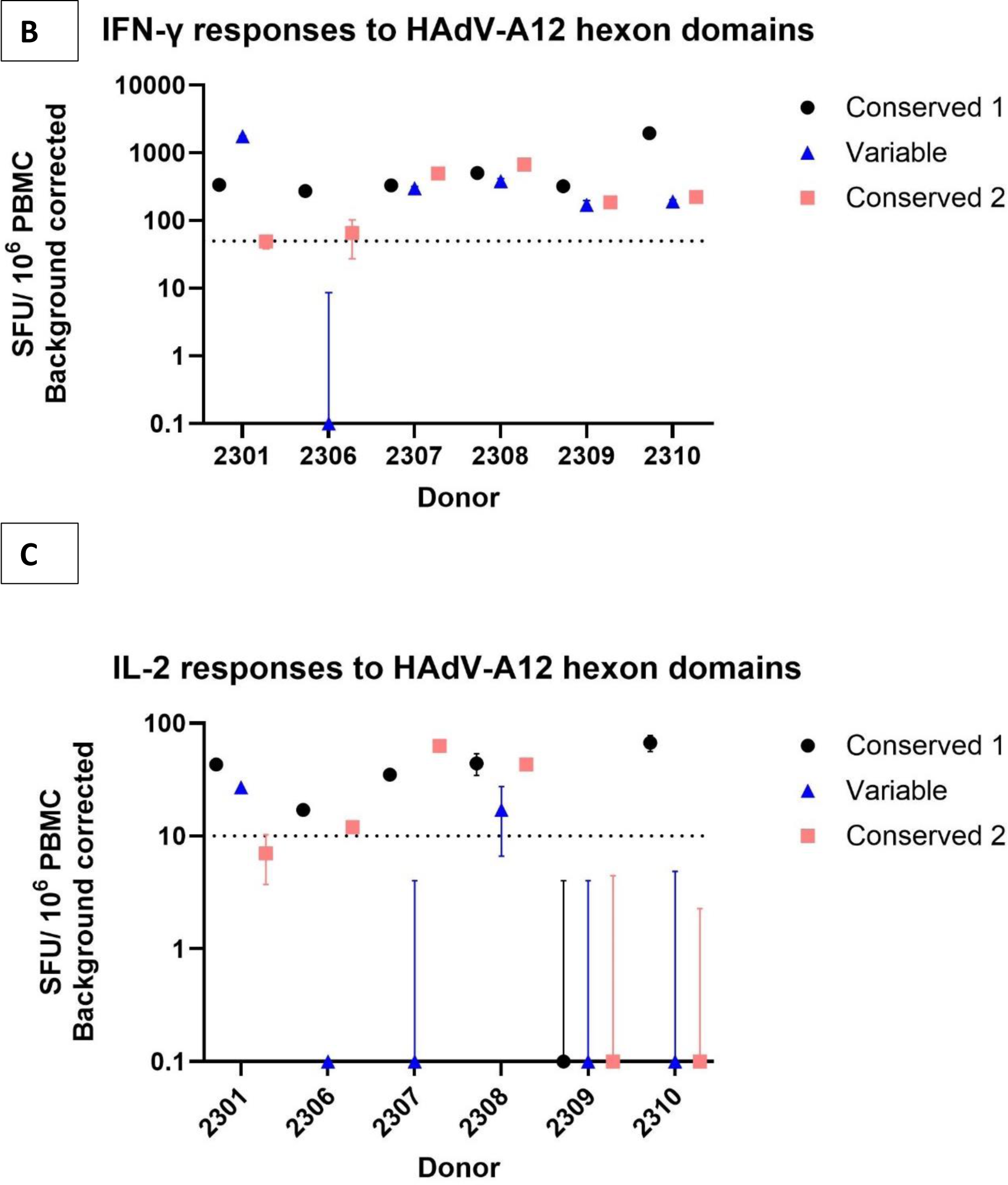
A: Diagram showing the location of the adenovirus A12 hexon in genomic context, and the composition of the conserved and variable peptide pools derived from this protein. B: Frequency of PBMC IFNγ responses to the HAdV-A12 hexon, divided into two conserved and one variable epitope pool. Points show the mean (n=3) and SEM for each donor and pool. The dotted line indicates a positive IFNγ response was defined as greater than 50 SFU/10^6 PBMC (background corrected). C: Frequency of PBMC IL2 responses to the conserved and variable epitope pools. The dotted line indicates a positive IL2 response was defined as greater than 10 SFU/10^6 PBMC (background corrected).

## Conclusions

We find that adenovirus-specific cellular immune responses to five HAdV species are widespread in UK blood donors, and include inflammatory cytokine responses to a widely-deployed SARS-CoV-2 vaccine vector backbone (ChAd-Y25, used in the development of the ChAdOx1 vaccine). Responses to the penton protein are less commonly detected, and at a lower frequency. We find that IFNγ responses to variable regions of the hexon protein may be more common than previously thought, particularly for genotype A12, while IL2 responses are often focused on conserved domains. We also present evidence that cross-type and type-specific IFNγ, but not IL2, responses have been boosted in ChAdOx1 recipients, with unknown consequences for population-level immunity or future adenovirus evolution.

## Data Availability

All data produced in the present study are available upon reasonable request to the authors and will be made available in supplementary material upon publication.

## Abbreviations

ChAd: Chimpanzee adenovirus
ELISA: Enzyme-linked immunosorbent assay
FluoroSpot: Fluorescence-linked immunosorbent spot
HAdV: Human adenovirus
HRA: Health Research Authority
IFNγ: Interferon gamma
IL2: Interleukin 2
nAbs: Neutralising antibodies
NHSBT: National Health Service Blood & Transplant
PBMC: Peripheral blood mononuclear cells
SARS-CoV-2: Severe acute respiratory syndrome coronavirus 2
SFU: Spot forming units

## Acknowledgements

The authors thank Eleanor Lim, Mark Wills and Marina Metaxaki for phlebotomy support and James V. Taylor for Matlab assistance. This research was supported by the Cambridge NIHR BRC Cell Phenotyping Hub.

## Author contributions

CJH conceived and designed the study. CJH, BJR and CNA supervised the study. RM, AWL, JPH, BACK and CJH performed the experiments and analysed the data. JPH, CNA, BACK, and CJH wrote the manuscript. All authors commented on the manuscript and approved submission.

## References

1 Benko M, Aoki K, Arnberg N, Davison AJ, Echavarria M, Hess M et al. ICTV Virus Taxonomy Profile: Adenoviridae 2022. Journal of General Virology 2022; 103: 001721.

2 Lion T, Wold W. Adenoviruses. In: Damania B, Cohen J (eds). Fields Virology: DNA Viruses. Wolters Kluwer, 2021, pp 129–171.

3 Al-Heeti OM, Cathro HP, Ison MG. Adenovirus Infection and Transplantation. Transplantation 2022; 106: 920–927.

4 Gray GC, Erdman DD. Adenovirus Vaccines. In: Plotkin’s Vaccines. Elsevier, 2018, pp 121–133.e8.

5 Walsh MP, Chintakuntlawar A, Robinson CM, Madisch I, Harrach B, Hudson NR et al. Evidence of Molecular Evolution Driven by Recombination Events Influencing Tropism in a Novel Human Adenovirus that Causes Epidemic Keratoconjunctivitis. PLoS One 2009; 4: e5635.

6 Duerr A, Huang Y, Buchbinder S, Coombs RW, Sanchez J, Del Rio C et al. Extended Follow-up Confirms Early Vaccine-Enhanced Risk of HIV Acquisition and Demonstrates Waning Effect Over Time Among Participants in a Randomized Trial of Recombinant Adenovirus HIV Vaccine (Step Study). J Infect Dis 2012; 206: 258–266.

7 Sumida SM, Truitt DM, Kishko MG, Arthur JC, Jackson SS, Gorgone DA et al. Neutralizing Antibodies and CD8 + T Lymphocytes both Contribute to Immunity to Adenovirus Serotype 5 Vaccine Vectors. J Virol 2004; 78: 2666–2673.

8 Walti CS, Stuehler C, Palianina D, Khanna N. Immunocompromised host section: Adoptive T-cell therapy for dsDNA viruses in allogeneic hematopoietic cell transplant recipients. Curr Opin Infect Dis 2022; 35: 302–311.

9 Hutnick NA, Carnathan D, Demers K, Makedonas G, Ertl HCJ, Betts MR. Adenovirus-specific human T cells are pervasive, polyfunctional, and cross-reactive. Vaccine 2010; 28: 1932–1941.

10 Leen AM, Sili U, Vanin EF, Jewell AM, Xie W, Vignali D et al. Conserved CTL epitopes on the adenovirus hexon protein expand subgroup cross-reactive and subgroup-specific CD8+ T cells. Blood 2004; 104: 2432–2440.

11 Veltrop-Duits LA, Heemskerk B, Sombroek CC, van Vreeswijk T, Gubbels S, Toes REM et al. Human CD4+ T cells stimulated by conserved adenovirus 5 hexon peptides recognize cells infected with different species of human adenovirus. Eur J Immunol 2006; 36: 2410–2423.

12 Leen AM, Christin A, Khalil M, Weiss H, Gee AP, Brenner MK et al. Identification of Hexon-Specific CD4 and CD8 T-Cell Epitopes for Vaccine and Immunotherapy. J Virol 2008; 82: 546–554.

13 Tang J, Olive M, Pulmanausahakul R, Schnell M, Flomenberg N, Eisenlohr L et al. Human CD8+ cytotoxic T cell responses to adenovirus capsid proteins. Virology 2006; 350: 312–322.

14 Toogood CIA, Crompton J, Hay RT. Antipeptide antisera define neutralizing epitopes on the adenovirus hexon. Journal of General Virology 1992; 73: 1429–1435.

15 Pichla-Gollon SL, Drinker M, Zhou X, Xue F, Rux JJ, Gao G-P et al. Structure-Based Identification of a Major Neutralizing Site in an Adenovirus Hexon. J Virol 2007; 81: 1680–1689.

16 Roberts DM, Nanda A, Havenga MJE, Abbink P, Lynch DM, Ewald BA et al. Hexon-chimaeric adenovirus serotype 5 vectors circumvent pre-existing anti-vector immunity. Nature 2006 441:7090 2006; 441: 239–243.

17 MHRA FOI Team. AstraZeneca COVID-19 vaccination first doses given FOI 22/1217. 2023.

18 Radukic MT, To Le D, Müller KM. Nucleic Acid Sequence Composition of the Oxford – AstraZeneca Vaccine ChAdOx1 nCoV-19 (AZD1222, Vaxzevria). 2021. doi:10.21203/RS.3.RS-799338/V1.

19 Dicks MDJ, Spencer AJ, Edwards NJ, Wadell G, Bojang K, Gilbert SC et al. A Novel Chimpanzee Adenovirus Vector with Low Human Seroprevalence: Improved Systems for Vector Derivation and Comparative Immunogenicity. PLoS One 2012; 7: e40385.

20 Poole E, Groves I, Jackson S, Wills M, Sinclair J. Using Primary Human Cells to Analyze Human Cytomegalovirus Biology. Methods Mol Biol 2021; 2244: 51–81.

21 Houldcroft CJ, Jackson SE, Lim EY, Sedikides GX, Davies EL, Atkinson C et al. Assessing Anti-HCMV Cell Mediated Immune Responses in Transplant Recipients and Healthy Controls Using a Novel Functional Assay. Front Cell Infect Microbiol 2020; 10: 275.

22 Krishna BA, Lim EY, Mactavous L, Lyons PA, Doffinger R, Bradley JR et al. Evidence of previous SARS-CoV-2 infection in seronegative patients with long COVID. EBioMedicine 2022; 81. doi:10.1016/J.EBIOM.2022.104129.

23 Koukoulias K, Papayanni PG, Jones J, Kuvalekar M, Watanabe A, Velazquez Y et al. Assessment of the cytolytic potential of a multivirus-targeted T cell therapy using a vital dye-based, flow cytometric assay. Front Immunol 2023; 14: 1299512.

24 Barnes E, Folgori A, Capone S, Swadling L, Aston S, Kurioka A et al. Novel adenovirus-based vaccines induce broad and sustained T cell responses to HCV in man. Sci Transl Med 2012; 4. doi:10.1126/SCITRANSLMED.3003155/SUPPL_FILE/4-115RA1_SM.PDF.

25 Jones DL. Fathom Toolbox for Matlab | USF College of Marine Science. 2017.https://www.usf.edu/marine-science/research/matlab-resources/fathom-toolbox-for-matlab.aspx (accessed 15 Apr2024).

26 Mennechet FJD, Paris O, Ouoba AR, Salazar Arenas S, Sirima SB, Takoudjou Dzomo GR et al. A review of 65 years of human adenovirus seroprevalence. Expert Rev Vaccines 2019; 18: 597–613.

27 Klann PJ, Wang X, Elfert A, Zhang W, Köhler C, Güttsches AK et al. Seroprevalence of Binding and Neutralizing Antibodies against 39 Human Adenovirus Types in Patients with Neuromuscular Disorders. Viruses 2023; 15: 79.

28 Wang X, Kerkmann L, Hetzel M, Windmann S, Trilling M, Zhang W et al. Analysis of the Prevalence of Binding and Neutralizing Antibodies against 39 Human Adenovirus Types in Student Cohorts Reveals Low-Prevalence Types and a Decline in Binding Antibody Levels during the SARS-CoV-2 Pandemic. J Virol 2022; 96. doi:10.1128/JVI.01133-22/ASSET/690BA172-2893-41C1-A330-92BF27AE837F/ASSETS/IMAGES/LARGE/JVI.01133-22-F006.JPG.

29 Slifka MK, Whitton JL. Antigen-Specific Regulation of T Cell–Mediated Cytokine Production. Immunity 2000; 12: 451–457.

30 Tischer S, Geyeregger R, Kwoczek J, Heim A, Figueiredo C, Blasczyk R et al. Discovery of immunodominant T-cell epitopes reveals penton protein as a second immunodominant target in human adenovirus infection. J Transl Med 2016; 14: 1–16.

31 Tori ME, Chontos-Komorowski J, Stacy J, Lamson DM, George KS, Lail AT et al. Identification of Large Adenovirus Infection Outbreak at University by Multipathogen Testing, South Carolina, USA, 2022 - Volume 30, Number 2—February 2024 - Emerging Infectious Diseases journal - CDC. Emerg Infect Dis 2024; 30: 358–362.

32 Smith CA, Woodruff LS, Rooney C, Kitchingman GR. Extensive Cross-Reactivity of Adenovirus-Specific Cytotoxic T Cells. https://home.liebertpub.com/hum 2008; 9: 1419–1427.

33 Onion D, Crompton LJ, Milligan DW, Moss PAH, Lee SP, Mautner V. The CD4+ T-cell response to adenovirus is focused against conserved residues within the hexon protein. Journal of General Virology 2007; 88: 2417–2425.

34 Feuchtinger T, Matthes-Martin S, Richard C, Lion T, Fuhrer M, Hamprecht K et al. Safe adoptive transfer of virus-specific T-cell immunity for the treatment of systemic adenovirus infection after allogeneic stem cell transplantation. Br J Haematol 2006; 134: 64–76.

35 Frahm N, DeCamp AC, Friedrich DP, Carter DK, Defawe OD, Kublin JG et al. Human adenovirus-specific T cells modulate HIV-specific T cell responses to an Ad5-vectored HIV-1 vaccine. J Clin Invest 2012; 122: 359–367.

36 Becken BA, Lamson DM, Gonzalez G, Patel S, St George K, Kajon AE. A Fulminant Case of Adenovirus Genotype C108 Infection in a Pediatric Stem Cell Transplant Recipient with x-Linked Lymphoproliferative Syndrome Type 1. Viruses 2024, Vol 16, Page 137 2024; 16: 137.

37 Al-Akioui Sanz K, Echecopar Parente C, Ferreras C, Menéndez Ribes M, Navarro A, Mestre C et al. Familial CD45RA– T cells to treat severe refractory infections in immunocompromised patients. Front Med (Lausanne) 2023; 10: 1083215.

38 Cervantes-Torres J, Cabello-Gutiérrez C, Dolores ·, Ayón-Núñez A, Soldevila G, Olguin-Alor R et al. Caveats of chimpanzee ChAdOx1 adenovirus-vectored vaccines to boost anti-SARS-CoV-2 protective immunity in mice. Applied Microbiology and Biotechnology 2024 108:1 2024; 108: 1–15.

39 Amosova IV, Timoshicheva TA, Kadyrova RA, Zabrodskaya YA, Vakin VS, Grudinin MP et al. The investigation of the dynamics of changes in neutralizing antibody titers against type 5 adenovirus in the context of vaccination against a new coronavirus infection. Virology 2024; 594: 110051.

40 Boyman O, Sprent J. The role of interleukin-2 during homeostasis and activation of the immune system. Nature Reviews Immunology 2012 12:3 2012; 12: 180–190.

